## Supplementary Materials for "Breakpoint modelling of temporal associations between non-pharmaceutical interventions and the incidence of symptomatic COVID-19 in the Republic of Ireland"

| Table S1. Pobal HP deprivation index components and descriptions | | |
| --- | --- | --- |
| Component Label | **Component Name** | **Component Description** |
| HPabs | Absolute HP Index Score | Composite measure of deprivation calculated for each SA, measured on a single scale across all census periods |
| HPrel | Relative HP Index Score | A measure of the level of deprivation in each SA relative to all other small areas surveyed |
| TOTPOP | Total population | Total population in each SA during each census period |
| POPCHG | Population Change | Percentage increase in population over the previous five years |
| AGEDEP | Age dependency rate | Percentage of population aged under 15 or over 64 years of age |
| LONEPA | Lone parent ratio | Percentage of households with children aged under 15 years and headed by a single parent |
| EDLOW | Primary education | Percentage of people in each SA with primary education as their highest level of education attainment |
| EDHIGH | Third level education | Percentage of people in each SA with third level education as their highest level of education attainment |
| HLPROF | Higher and lower professionals | Percentage of households headed by professionals or managerial and technical employees, including farmers with 100 acres or more |
| LSKILL | Proportion of semi-skilled and unskilled manual workers | Percentage of households in each SA headed by semi‐skilled or unskilled manual workers, including farmers with less than 30 acres |
| UNEMPM | Male unemployment rate | Rate of male unemployment in each SA |
| UNEMPF | Female unemployment rate | Rate of female unemployment in each SA |
| PEROOM | Persons per room | Mean number of persons per household room in each small area |
| LARENT | Local authority housing | Percentage of local authority housing in each SA |
| PRRENT | Privately rented housing | Percentage of privately rented housing in each SA |
| OHOUSE | Own home | Percentage of privately owned housing in each SA |

| **Table S2. Pobal HP Index absolute and relative deprivation index score classification** | |
| --- | --- |
| **Index Score** | **Level of deprivation/affluence** |
| **-40 to - 30** | Extremely disadvantaged |
| **-30 to -20** | Very disadvantaged |
| **-20 to -10** | Disadvantaged |
| **-10 to 0** | Marginally below average |
| **0 to 10** | Marginally above average |
| **10 to 20** | Affluent |
| **20 to 30** | Very affluent |
| **30 to 40** | Extremely affluent |

*Note.* Reprinted from the 2016 Pobal HP Deprivation Index for Small Areas

(SA): Introduction and Reference Tables, Trutz Haase Jonathan Pratschke,

September 2017
